# Modelling the impact of COVID-19 in Australia to inform transmission reducing measures and health system preparedness

**DOI:** 10.1101/2020.04.07.20056184

**Authors:** R Moss, J Wood, D Brown, F Shearer, AJ Black, AC Cheng, JM McCaw, J McVernon

**Affiliations:** Modelling and Simulation Unit, Melbourne School of Population and Global Health, The University of Melbourne; School of Public Health and Community Medicine, UNSW Sydney; Victorian Infectious Diseases Laboratory Epidemiology Unit at The Peter Doherty Institute for Infection and Immunity; The University of Melbourne and Royal Melbourne Hospital, Melbourne, Australia; School of Mathematical Sciences, University of Adelaide; Infection Prevention and Healthcare Epidemiology Unit, Alfred Health; School of Public Health and Preventive Medicine, Monash University; School of Mathematics and Statistics, The University of Melbourne; Infection and Immunity Theme, Murdoch Children’s Research Institute, the Royal Children’s Hospital, Melbourne

## Abstract

**Background:** The ability of global health systems to cope with increasing numbers of COVID-19 cases is of major concern. In readiness for this challenge, Australia has drawn on clinical pathway models developed over many years in preparation for influenza pandemics. These models have been used to estimate health care requirements for COVID-19 patients, in the context of broader public health measures.

**Methods:** An age and risk stratified transmission model of COVID-19 infection was used to simulate an unmitigated epidemic with parameter ranges reflecting uncertainty in current estimates of transmissibility and severity. Overlaid public health measures included case isolation and quarantine of contacts, and broadly applied social distancing. Clinical presentations and patient flows through the Australian health care system were simulated, including expansion of available intensive care capacity and alternative clinical assessment pathways.

**Findings:** An unmitigated COVID-19 epidemic would dramatically exceed the capacity of the Australian health system, over a prolonged period. Case isolation and contact quarantine alone will be insufficient to constrain case presentations within a feasible level of expansion of health sector capacity. Overlaid social restrictions will need to be applied at some level over the course of the epidemic to ensure that systems do not become overwhelmed, and that essential health sector functions, including care of COVID-19 patients, can be maintained. Attention to the full pathway of clinical care is needed to ensure access to critical care.

**Interpretation:** Reducing COVID-19 morbidity and mortality will rely on a combination of measures to strengthen and extend public health and clinical capacity, along with reduction of overall infection transmission in the community. Ongoing attention to maintaining and strengthening the capacity of health care systems and workers to manage cases is needed.

**Funding:** Australian Government Department of Health Office of Health Protection, Australian Government National Health and Medical Research Council

## Background

As of early April 2020, more than a million confirmed cases of COVID-19 have been reported worldwide, involving all global regions and resulting in over 50,000 deaths (1). Although the majority of cases are clinically mild or asymptomatic, early reports from China estimated that 20% of all COVID-19 patients progressed to severe disease and required hospitalisation, with 5-16% of these individuals going on to require management within an Intensive Care Unit (ICU) (2). Pulmonary disease leading to respiratory failure has been the major cause of mortality in severe cases (3).

The ability of health systems around the world to cope with increasing case numbers in coming months is of major concern. All levels of the system will be challenged, from primary care, pre-hospital and emergency department (ED) services, to inpatient units and ultimately ICUs. Stresses on clinical care provision will result in increased morbidity and mortality (4). These consequences have tragically already been observed even in high income countries that provide whole population access to quality medical care. Greater impacts will be observed over coming months in low and middle-income countries where access to high level care is extremely limited. Availability of ICU beds and ventilators has proven critical for the adequate management of severe cases, with overwhelming demand initiating complex ethical discussions about rationing of scarce resources (5).

In readiness for this challenge, Australia has drawn on approaches developed over many years to prepare for influenza pandemics (6) and rapidly produced a national COVID-19 pandemic plan (7). The plan has reoriented relevant strategies towards this new pathogen, based on emerging understanding of its anticipated transmissibility and severity, which are the determinants of clinical impact (8). Early imposition of stringent border measures, high levels of testing, active case-finding and quarantine of contacts have all bought time to reinforce public health and clinical capacity. However, a recent influx of cases among travelers returning from countries with rapidly growing epidemics has been associated with establishment of community transmission in several Australian states. As of 31^st^ March 2020, 4,359 cases had been reported, with 18 deaths (9).

This study reports on the use of a clinical care pathways model that represents the capacity of the Australian health system. This framework was initially developed for influenza pandemic preparedness (10), and has been modified to estimate health care requirements for COVID-19 patients and inform needed service expansion. The ability of different sectors to meet anticipated demand was assessed by modelling plausible COVID-19 epidemic scenarios, overlaid on available capacity and models of patient flow and care delivery. An unmitigated outbreak is anticipated to completely overwhelm the system. Given realistic limits on capacity expansion, these models have made the case for ongoing case targeted measures, combined with broader social restrictions, to reduce transmission and ‘flatten the curve’ of the local epidemic to preserve health sector continuity.

## Methods

### Disease transmission model

We developed an age and risk stratified transmission model of COVID-19 infection based on a susceptible-exposed-infected-recovered (SEIR) paradigm, described in full in supplementary material. Transmission parameters were based on information synthesis from multiple sources, with an assumed *R*_*0*_ of 2.53, and a doubling time of 6.4 days (Table 1). Potential for pre-symptomatic transmission was assumed within 48 hours prior to symptom onset. While there is an increasing body of evidence regarding requirements of hospitalised patients for critical care, considerable uncertainty remains regarding the full ‘pyramid’ of mild and moderately symptomatic disease. We therefore simulated a range of scenarios using Latin Hypercube Sampling from distributions in which the proportion of all infections severe enough to require hospitalisation ranged between 4.3 and 8.6%. These totals represent the aggregate of strongly age-skewed parameter assumptions (Table 2). For each scenario, corresponding distributions of mild cases presenting to primary care were sampled, ranging from 30-45% at the lower range of the ‘severe’ spectrum, to 50-75% for the most extreme case (increasing linearly between the two). Cases not presenting to the health system were assumed ‘undetected’ without differentiation between those with mild or no symptoms.

**Table 1:**
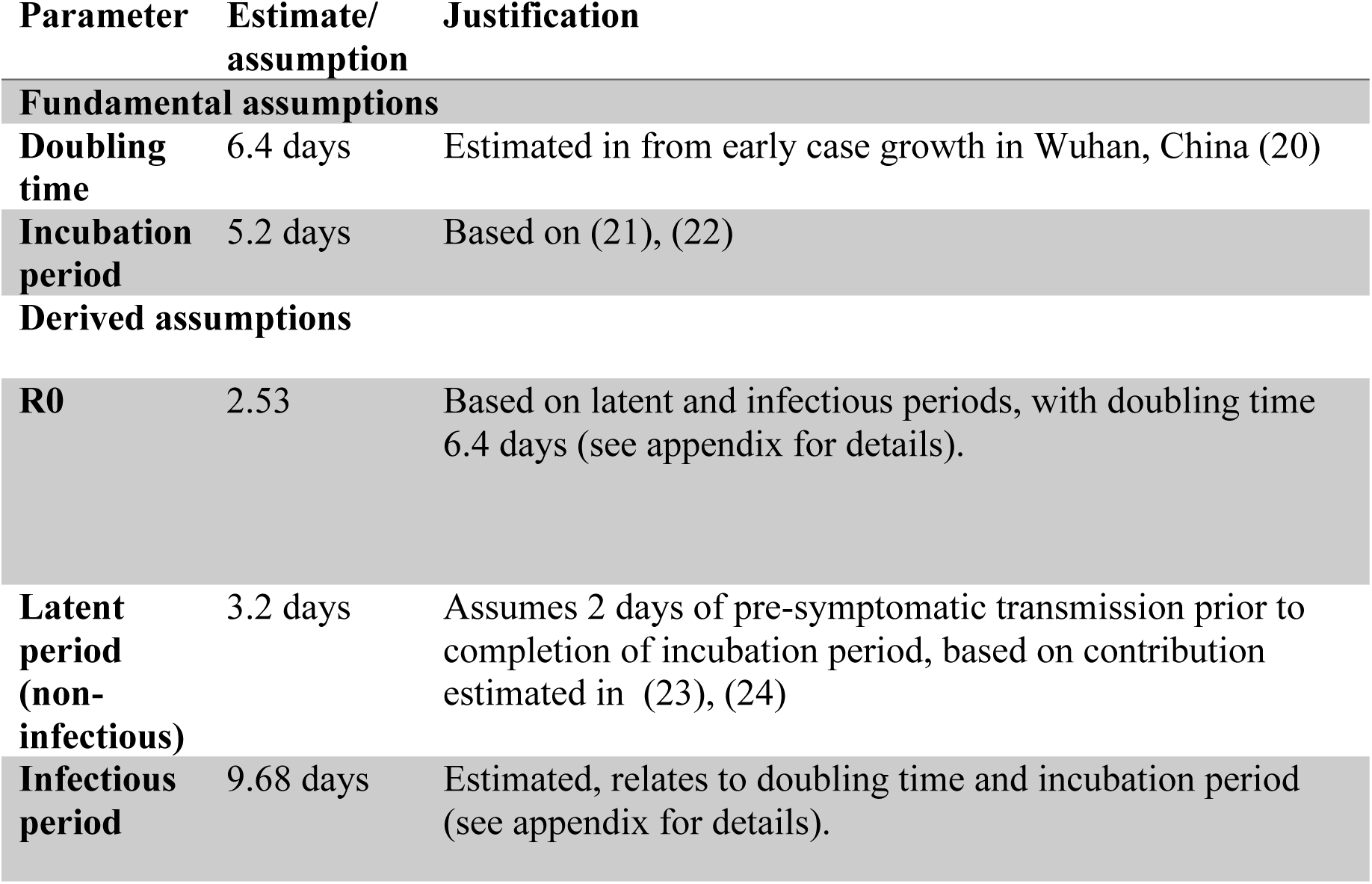
COVID-19 model transmission parameter assumptions.

| Parameter | Estimate/<br>assumption | Justification |
| --- | --- | --- |
| **Fundamental assumptions** |  |  |
| **Doubling time** | 6.4 days | Estimated in from early case growth in Wuhan, China (20) |
| **Incubation period** | 5.2 days | Based on (21), (22) |
| **Derived assumptions** |  |  |
| **R0** | 2.53 | Based on latent and infectious periods, with doubling time 6.4 days (see appendix for details). |
| **Latent period (non-infectious)** | 3.2 days | Assumes 2 days of pre-symptomatic transmission prior to completion of incubation period, based on contribution estimated in (23), (24) |
| **Infectious period** | 9.68 days | Estimated, relates to doubling time and incubation period (see appendix for details). |

**Table 2:** COVID-19 model severity parameter assumptions, relative to all denominator infections.

| Age group (years) | %Hospitalised Range<sup>s</sup> | %Require ICU<sup>^</sup> |
| --- | --- | --- |
| 0-9 | 0.03% - 0.06% | 0.01% - 0.02% |
| 10-19 | 0.03% - 0.06% | 0.01% - 0.02% |
| 20-29 | 0.39% - 0.78% | 0.11% - 0.23% |
| 30-39 | 1.45% - 2.90% | 0.43% - 0.85% |
| 40-49 | 2.55% - 5.11% | 0.75% - 1.50% |
| 50-59 | 4.95% - 9.90% | 1.45% - 2.91% |
| 60-69 | 7.75% - 15.49% | 2.27% - 4.55% |
| 70-79 | 17.88% - 35.76% | 5.25% - 10.50% |
| 80+ | 32.97% - 65.94% | 9.68% - 19.36% |
| **Mean bed days for inpatients<sup>&</sup>** |  |  |
| *Hospital* | 8 days |  |
| *ICU* | 10 days |  |

<sup>s</sup>Assumed proportional to ICU values and based on calibration to non-Hubei China severe case rates (see appendix for details).
<sup>^</sup>Combines use of data from (25), (26) and assumptions used in (13).
<sup>&</sup>Based on assumptions used in (13).

### Case targeted interventions

A case targeted public health intervention was simulated. Cases were isolated at the point of diagnosis (assumed 48 hours after symptom onset), limiting the effective infectious period and reducing infectiousness by 80%. Targeted quarantine of close contacts was implemented in the model framework by dynamic assignment of a transient ‘contact’ label. Each time a new infectious case appears in the model, a fixed number of temporary ‘contacts’ are labelled. Only contacts can progress through the exposed and infectious states, however the majority remain uninfected and return to their original ‘non-contact’ status within 72 hours. We assumed that 80% of identified contacts adhered to quarantine measures and that the overall infectiousness of truly exposed and infected contacts was halved by quarantine, given delayed and imperfect contact tracing and the risk of transmission to household members.

### Clinical pathways model

The model of patient flows for mild and severe disease is represented conceptually in Figure At baseline, we assume that half of available consulting and admission capacity across all sectors and services is available to COVID-19 patients. Mild cases present to primary care until capacity is exceeded. Severe cases access the hospital system through ED and from there are triaged to a ward or ICU bed according to need, if available. Requirements for critical care are assumed to increase steeply with age with the consequence that about 60% of all infections requiring ICU admission occur in individuals aged 70 years and over (Table 2). As ward beds reach capacity, the ability of emergency departments to triage patients is reduced because of bed block, meaning that not all patients who need care are medically assessed, although some will still be able to access primary care. The model allows for repeat representations within and between primary care and hospital services, and progression from ward to intensive care, with length of stay as shown (Table 2). Model structure and assumptions are based on publicly available data on the Australian health care system, and expert elicitation. Full model details are provided in Supplementary material.

### Critical care capacity expansion

The baseline assumption was that half of currently available ICU beds would be available to COVID-10 patients. Three capacity expansion scenarios were considered, assuming routine models of care for patient triage and assessment within the hospital system:

- Total ICU capacity expansion to 150% of baseline, doubling the number of beds available to treat COVID-19 patients (2 × ICU cap);
- Total ICU capacity expansion to 200% of baseline, tripling the number of beds available to treat COVID-19 patients (3 × ICU cap);
- Total ICU capacity expansion to 300% of baseline, increasing by fivefold the number of beds available to treat COVID-19 patients (5 × ICU cap).

A theoretical alternative clinical pathway with constraints on bed numbers but double the capacity to assess severe cases presenting to hospital was also considered (COVID-19 clinics). The purpose of including this pathway is to reveal unmet clinical need arising when bed block constrains ED triage capacity, potentially preventing needed admissions to ICU.

### Social distancing interventions

Broad based social distancing measures overcome ongoing opportunities for transmission arising from imperfect ascertainment of all cases and contacts, and from pre-symptomatic and asymptomatic individuals. In settings where non-pharmaceutical social interventions have been applied, associated case targeted measures have also been in place, making the effectiveness of each difficult to quantify (11). Data from Hong Kong showing a reduction in *influenza* incidence arising from a combination of distancing measures introduced in response to COVID-19 provides good evidence of generalised transmission reduction (12). However, the relative quantitative contributions of different interventions eg cancellation of mass gatherings, distance working, closure of schools, cessation of non-essential services, etc cannot be reliably differentiated at this time (13).

We therefore focused on the overall objective of distancing, which is to reduce the reproduction number. We modelled the impact of constraining spread by 25% and 33%, overlaid on existing case targeted interventions. This degree of suppression is consistent with observed impacts of combined measures less restrictive than total lockdown (13). These reductions in transmission equated to input reproduction numbers of 1.90 and 1.69, respectively, with the effective reproduction number in each scenario further reduced by quarantine and isolation measures, which limit spread of established infection.

## Findings

An unmitigated COVID-19 epidemic would dramatically exceed the capacity of the Australian health system, over a prolonged period (Figure 2). Case isolation and contact quarantine applied at the same level of effective coverage throughout the epidemic have the potential to substantially reduce transmission. By ‘flattening the curve’ they produce a prolonged epidemic with lower peak incidence (Figure 2) and fewer overall infections. Epidemic scenarios with higher assumed severity (95^th^ centile case) are more effectively delayed by these public health measures than less severe scenarios (50^th^ centile case). This finding is because a higher proportion of all infections present to health services and can be identified for isolation and contact tracing. In a mitigated epidemic, overall utilisation of the health system is increased, because more patients are able to access needed care over the extended epidemic duration (Supplementary Figure 1A).

**Figure 1:**
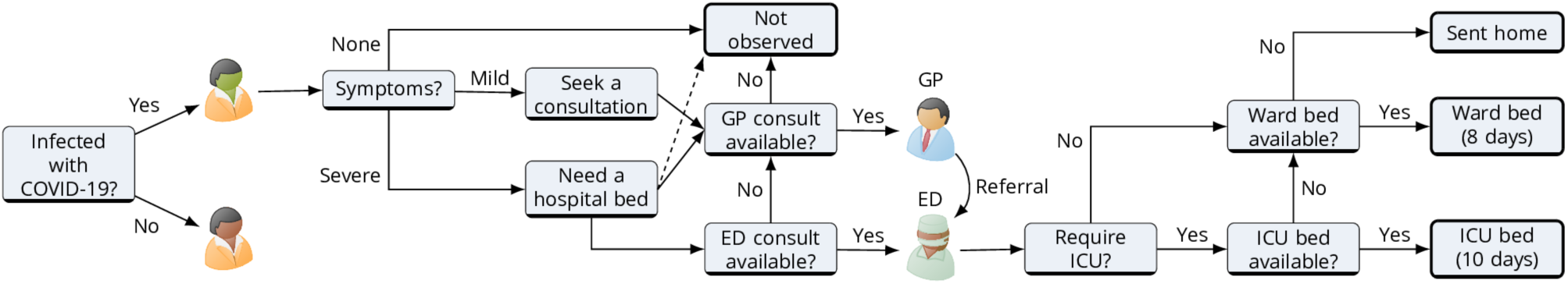
Representation of the clinical pathways model, that captures presentations for mild and severe illness, assuming that the former are managed within primary care.

**Figure 2:**
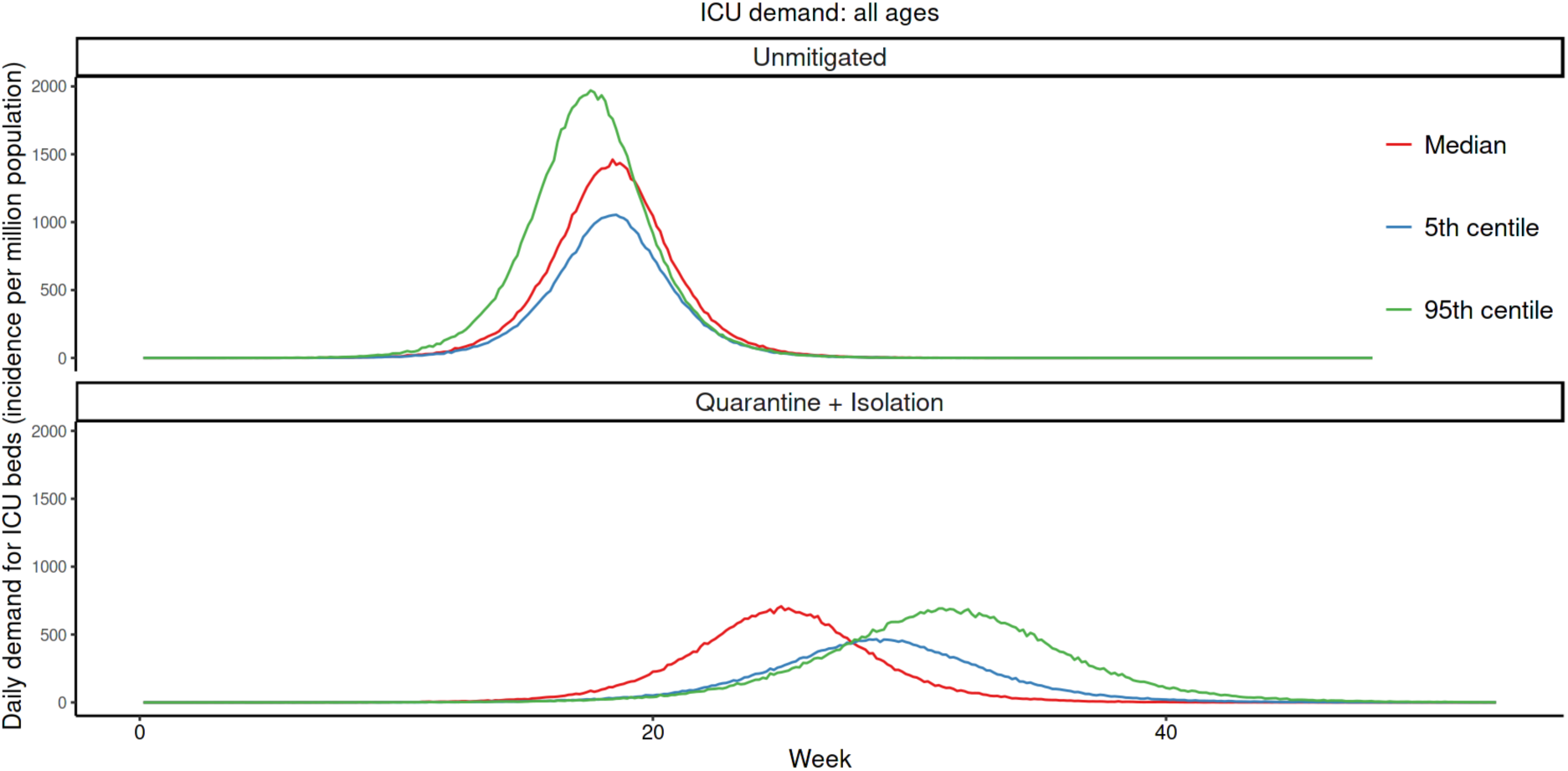
Daily incident ICU admission demand per million population in an unmitigated (upper series) COVID-19 epidemic, compared with one mitigated by case targeted public health measures (lower series). Lines represent single simulations based on median (red), 5^th^ (blue) or 95^th^ (green) centile parameter assumptions.

Increasing the number of ICU beds available to patients with COVID-19 reduces the period over which ICU capacity is anticipated to be exceeded, potentially by more than half (Figure 3A). The duration of exceedance for each capacity scenario is increased by quarantine and isolation because the overall epidemic is longer (Figure 3A). During the period of exceedance, a degree of unmet need remains, even for the mitigated scenario (Figure 3B). A five-fold increase in the number of ICU beds available to patients with COVID-19 dramatically reduces the period and peak of excess demand (Figures 3A, B).

**Figure 3A:**
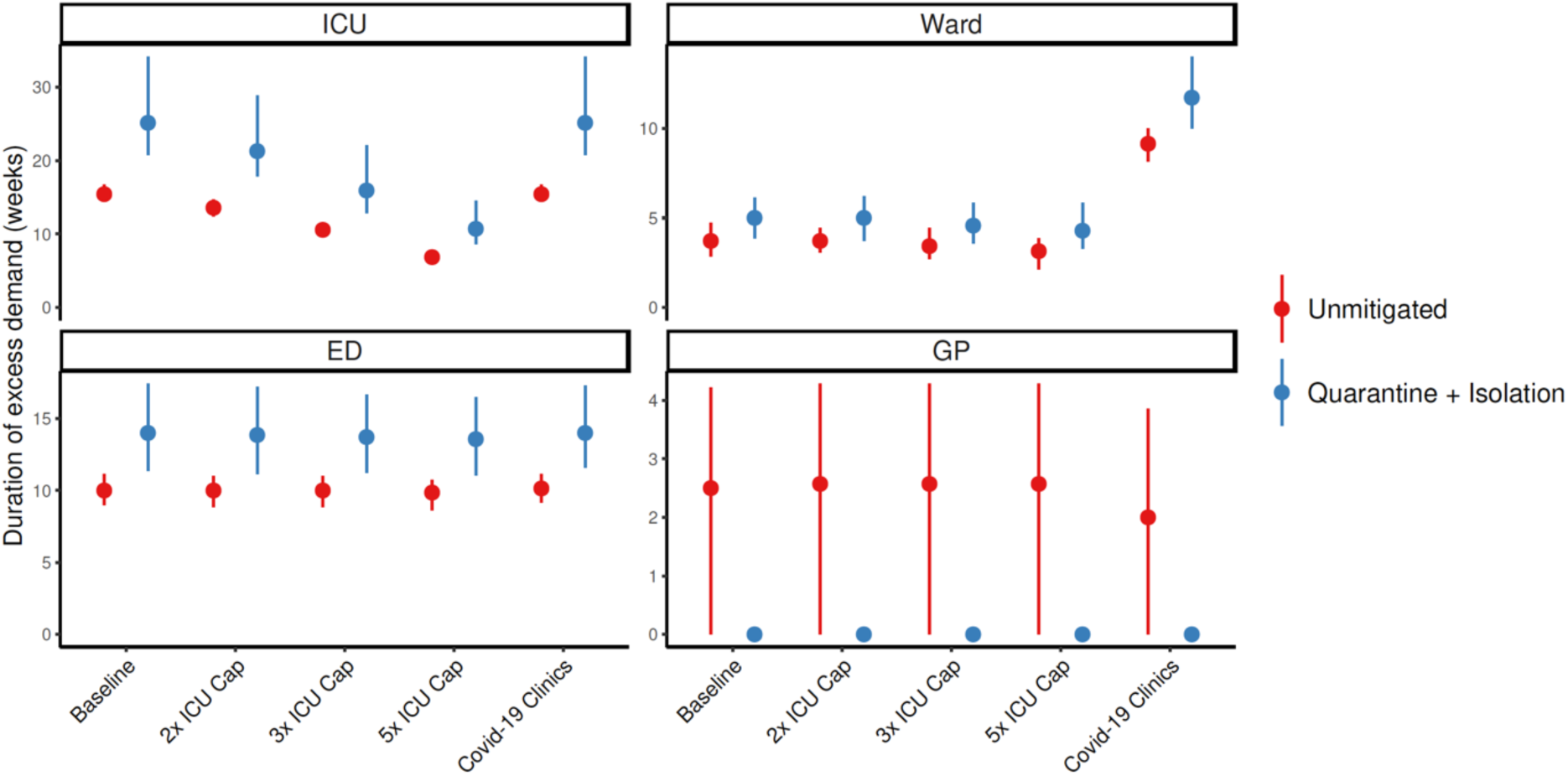
Duration of excess demand by sector over the course of the epidemic, by sector, for unmitigated (red) and mitigated (blue) scenarios. ICU capacity exceedance for COVID-19 admissions is compared for baseline, double, and nine times ICU capacity. The ‘COVID-19 clinics’ scenario reflects an alternative triage pathway, and baseline capacity. Dots denote medians, lines range from 5^th^ to 95^th^ centiles of simulations.

**Figure 3B:**
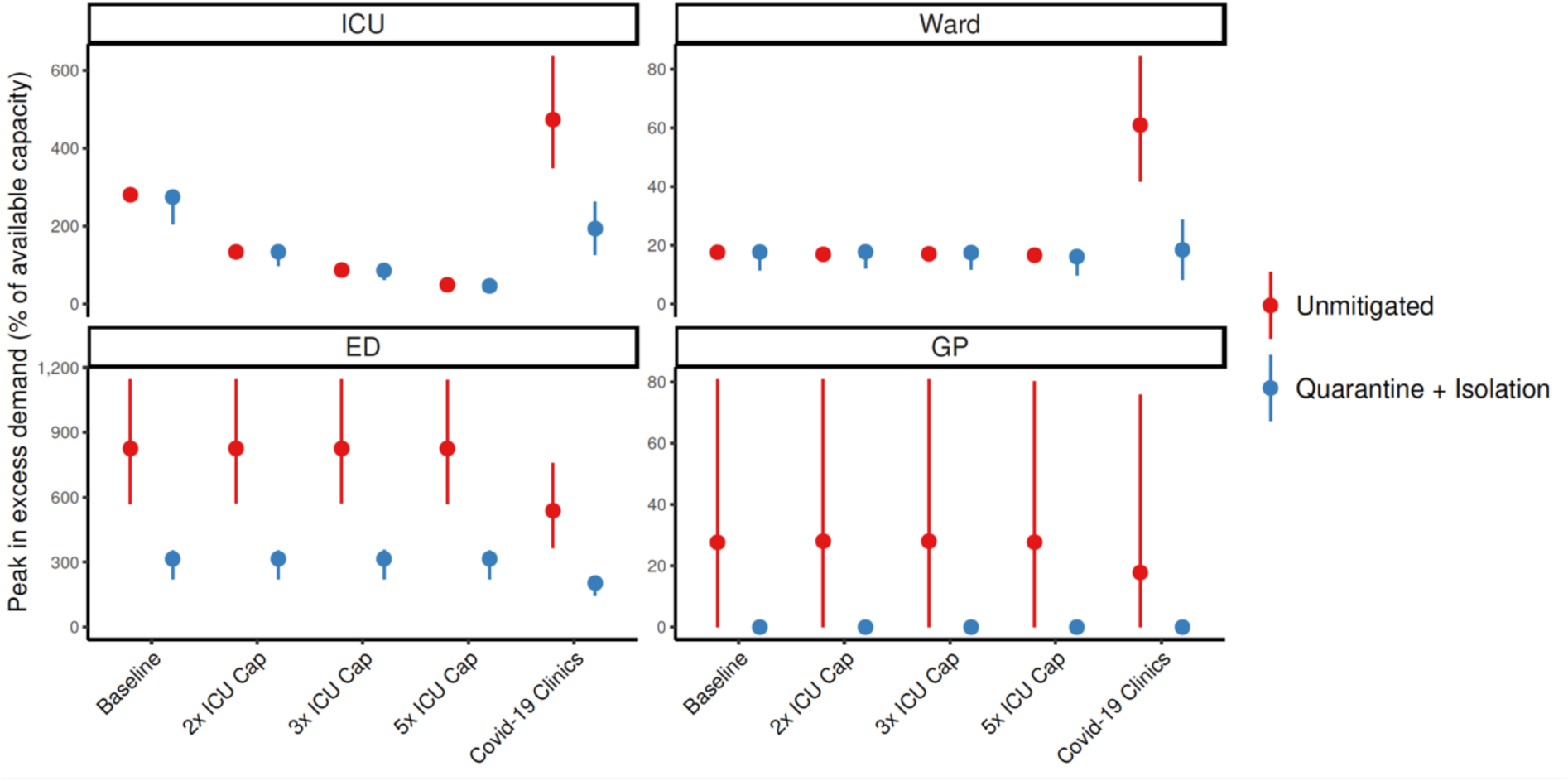
Peak excess demand by sector expressed as % available capacity, for unmitigated (red) and mitigated (blue) scenarios. This excess is compared for baseline, double, and nine times ICU capacity. The ‘COVID-19 clinics’ scenario reflects an alternative triage pathway, and baseline capacity. Dots denote medians, lines range from 5^th^ to 95^th^ centiles of simulations.

These figures do not accurately reflect the true requirement for services however, as blocks in assessment pathways resulting from emergency department and ward overload are an upstream constraint on incident ICU admissions. The alternative triage scenario (COVID-19 clinic) reveals a high level of unmet clinical need for both ward and critical care beds given baseline bed capacity (Figures 3A, 3B). Case targeted measures overcome this limitation to some extent, and effectively improve overall access to care (Figure 3A, 3B). Overall, if ICU beds available to COVID-19 patients are doubled, between 10 and 30% of those who require critical care receive it. This proportion rises to approximately 20-40% if capacity increases by five-fold (Supplementary Figure 1A). These figures are quantified as total excess demand per million over the course of the epidemic (Figure 1B).

As can be seen from these simulated scenarios, case isolation and contact quarantine alone will be insufficient to keep clinical requirements of COVID-19 cases within plausibly achievable expansion of health system capacity, even if very high and likely unrealistic levels of case finding can be maintained. We therefore explored the effects of additional social distancing measures that reduced input reproduction numbers of 25 and 33% on ICU requirements in relation to these same clinical care capacity constraints (Figure 4). Simulations assume ongoing application of measures of fixed effectiveness, which is also unlikely to be consistently achievable over an extended duration.

**Figure 4:**
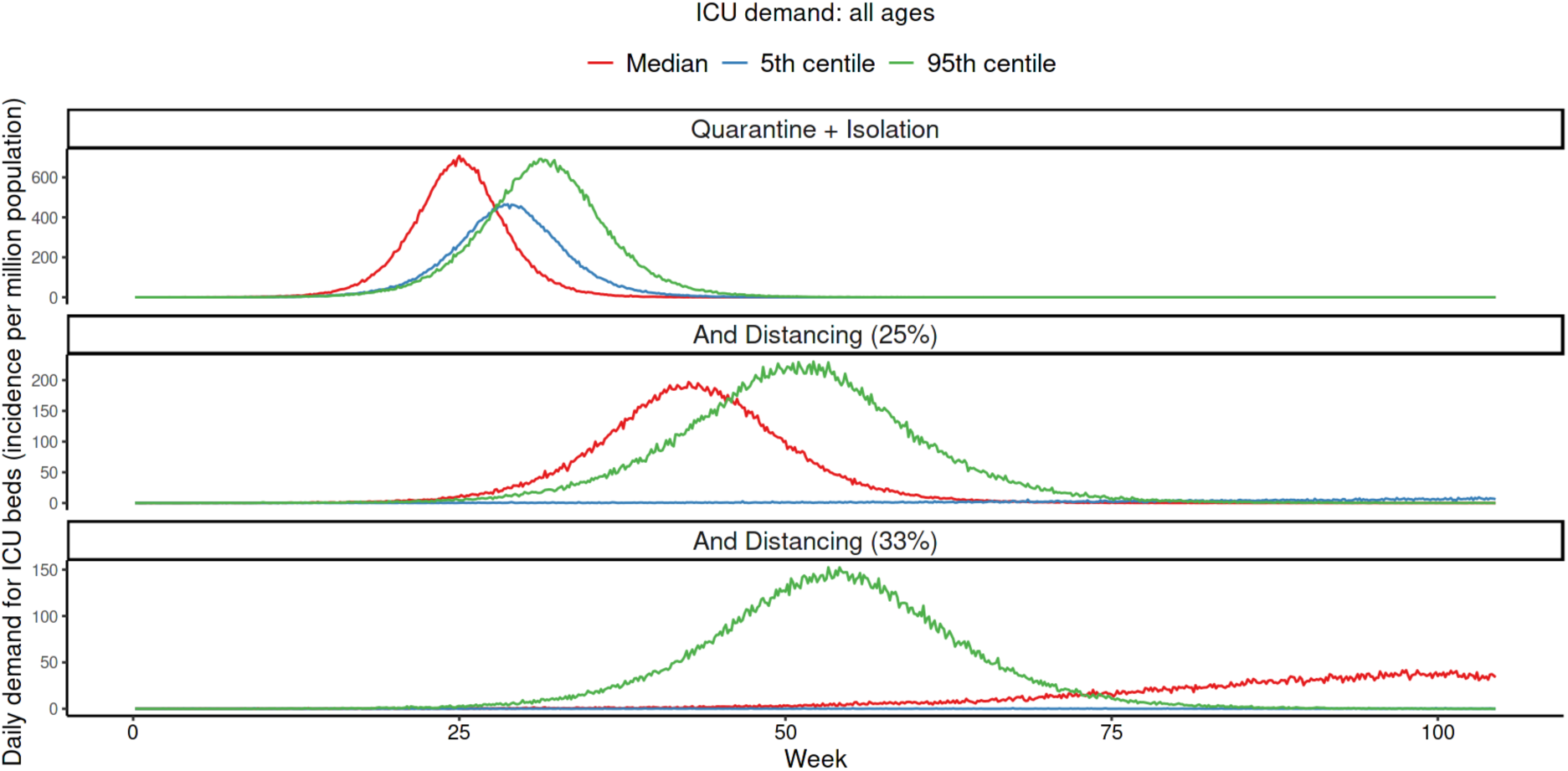
Daily incident ICU admission demand per million population comparing mitigation achievable with quarantine and isolation alone (upper series) with that achievable when 25% (middle series) or 33% (lower series) social distancing is overlaid. Lines represent single simulations based on median (red), 5^th^ (blue) or 95^th^ (green) centile parameter assumptions.

The overlay of distancing measures, applied from the initial stages of the epidemic and maintained throughout, suppresses epidemic growth to a level that is within the range of plausible ICU capacity expansion. The duration of ICU exceedance remains long in the 25% case (Figure 5A) but this overflow occurs to a far lesser degree than following case targeted strategies only (Figure 5B). As anticipated, a greater reduction in transmission (33%) achieves greater benefits. Importantly, pressure on ED consultations and ward beds is also substantially eased in these scenarios, maintaining capacity along the full pathway of care. As a result, the proportion of critical cases that can access care is greatly increased. Transmission reduction of 33% makes treatment of all cases achievable in the majority of simulations if three to five-fold ICU bed capacity can be achieved (Supplementary figure 1B). This improvement is reflected in a large reduction in unmet need (Supplementary figure 2B).

**Figure 5A:**
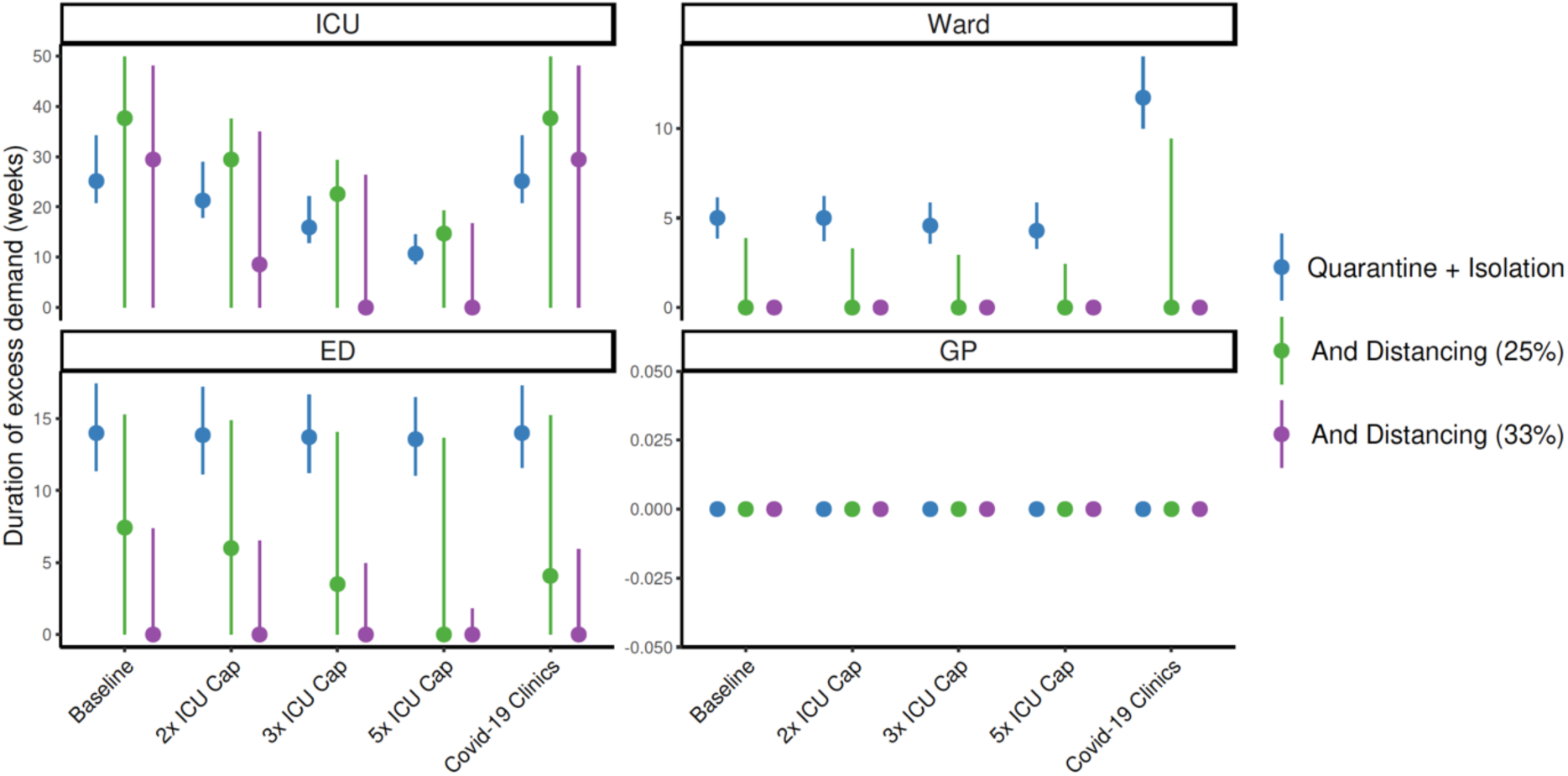
Duration of excess demand by sector over the course of the epidemic, by sector, for quarantine and isolation (blue) scenarios, with overlaid social distancing measures to reduce transmission by 25% (green) and 33% (purple). ICU capacity exceedance for COVID-19 admissions is compared for baseline, 2, 3 and 5 × ICU capacity. The ‘COVID-19 clinics’ scenario reflects an alternative triage pathway, and baseline capacity. Dots denote medians, lines range from 5^th^ to 95^th^ centiles of simulations.

**Figure 5B:**
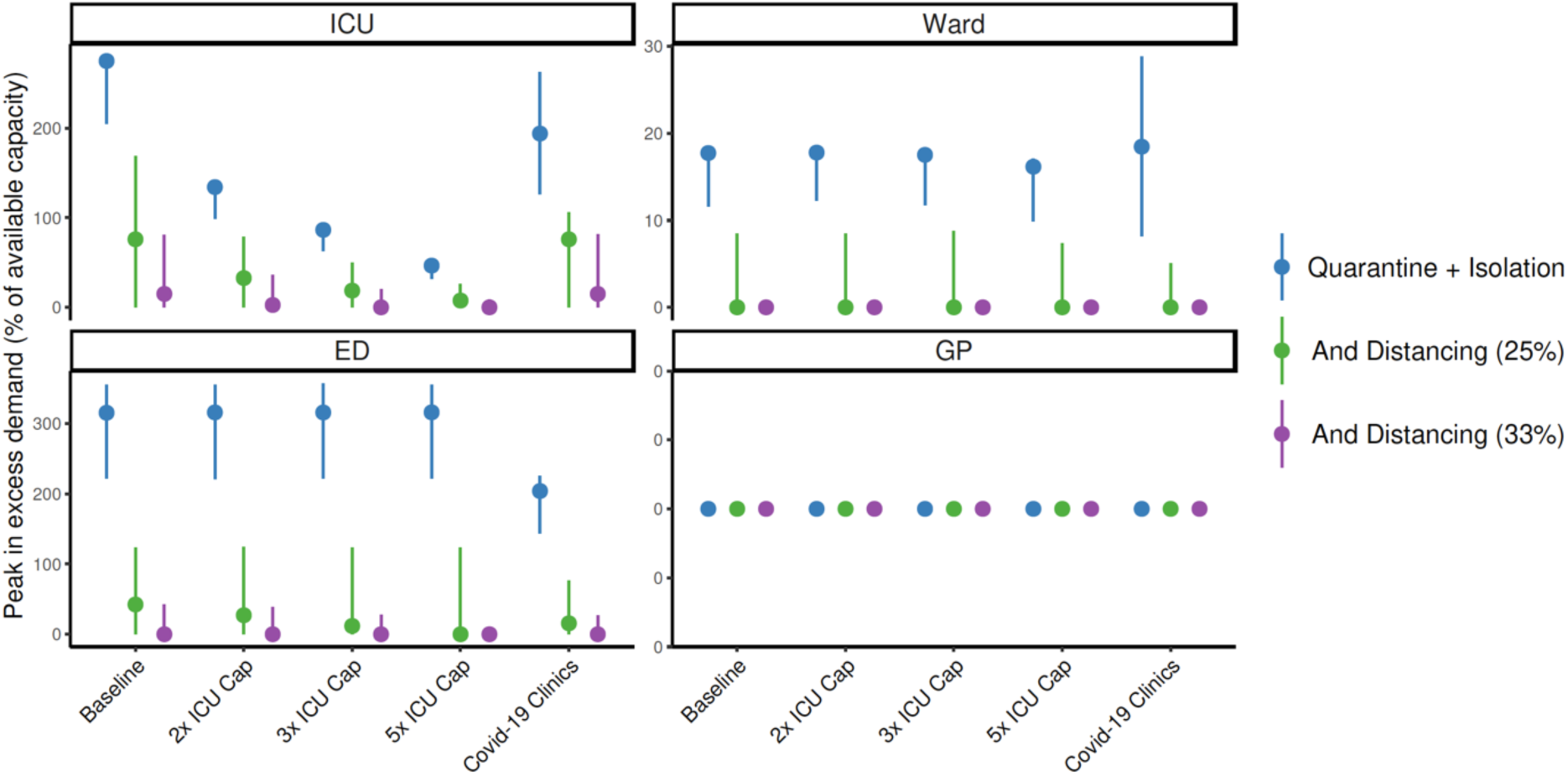
Peak excess demand by sector expressed as % available capacity, for quarantine and isolation (blue) scenarios, with overlaid social distancing measures to reduce transmission by 25% (green) and 33% (purple). This excess is compared for baseline, 2, 3, and 5 × ICU capacity. The ‘COVID-19 clinics’ scenario reflects an alternative triage pathway, and baseline capacity. Dots denote medians, lines range from 5^th^ to 95^th^ centiles of simulations.

## Interpretation

This modelling study has shown that an unmitigated COVID-19 epidemic would rapidly overwhelm Australia’s health sector capacity. Case targeted measures including isolation of those known to be infected, and quarantine of their close contacts, must remain an ongoing cornerstone of the public health response. While these interventions effectively reduce transmission, they are unlikely to be maintained at the high coverage modelled here throughout the epidemic course. As public health response capacity is exceeded, greater constraint of disease spread will be essential to ensure that feasible levels of expansion in available health care can maintain ongoing system functions, including care of COVID-19 patients. Broader based social and physical distancing measures reduce the number of potential contacts made by each case, minimising public health workload and supporting sustainable case targeted disease control efforts.

Our findings are consistent with a recently published model that relates the clinical burden of COVID-19 cases to global health sector capacity, characterised at high level. In unmitigated epidemics, demand rapidly outstrips supply, even in high income settings, by a factor of seven (14). As hospital bed capacity is strongly correlated with income, this factor is greatly increased in low and middle-income country settings, where underlying health status is also likely to be poorer (14). There is also marked variability globally in the definition of ‘intensive care’, even within high-income countries where the descriptor covers many levels of ventilatory and other support. The authors concur with our conclusion that social distancing measures to suppress disease are required to save lives. They also acknowledge that the marked social and economic consequences of such measures will limit their ongoing application, particularly in the settings where health systems are least able to cope with disease burden (14).

While much attention has been focused on expansion of available ICU beds per se, our clinical model reveals that critical care admissions are further limited by the ability to adequately assess patients during times of system stress. In line with model recommendations, Australia, with other countries, has implemented ‘COVID-19 clinics’ as an initial assessment pathway to reduce impacts on primary care and emergency department services (15). Such facilities have additional benefits of ensuring appropriate testing, in line with local case definitions, and reduce the overall consumption of personal protective equipment by cohorting likely infectious patients. Other measures to improve patient flows should also be considered based on evidence of bottlenecks as the epidemic progresses such as overflow expansion in EDs, encouraging and supporting home-based care, or early discharge to supported isolation facilities.

Quantitative findings from our model are limited by ongoing uncertainties about the true disease ‘pyramid’ for COVID-19, and a lack of nuanced information about determinants of severe disease, here represented by age as a best proxy. The clinical pathways model assumes that half of available bed capacity is available for patients with the disease but does not anticipate the seasonal surge in influenza admissions that may be overlaid with the epidemic peak, although even in our most recent severe season (2017) only 6% of hospital beds were occupied by influenza cases (16). Available beds will likely be increased by other factors such as secondary reductions in all respiratory infections and road trauma resulting from social restrictions, and purposive decisions to cancel non-essential surgery. Importantly, we do not consider health care worker absenteeism due to illness, carer responsibilities or burnout – all of which are anticipated challenges over a very prolonged epidemic accompanied by marked social disruption. We also cannot account for shortages in critical medical supplies as the true extent of these and their likely future impacts on service provision are presently unknown.

The model indicates that a combination of case-targeted and social measures will need to be applied over an extended period to reduce the rate of epidemic growth. In reality, it is likely that the stringency of imposed controls, their public acceptability and compliance will all vary over time. In Australia, compliance with isolation and self-quarantining was largely on the basis of trust in early response (February through March) but active monitoring and enforcement of these public health measures is now occurring in many jurisdictions. Hong Kong and Singapore initiated electronic monitoring technologies from the outset to track the location of individuals and enforce compliance (17). Proxy indicators of compliance such as transport and mobile phone data have informed understanding of the impact of social and movement restrictions on mobility and behaviour in other settings (11), and will be further investigated in the Australian context.

The effectiveness of multiple distancing measures including lockdown has been demonstrated in Europe, but the contributions of individual measures cannot yet be reliably differentiated (13). The impact of local measures to curb transmission will be estimated from real time data on epidemic growth in the Australian context, based on multiple epidemiological and clinical data streams. Estimates of the local effective reproduction number will enable forecasting of epidemic trajectories (18) to be fed into the analysis pathway presented here. Anticipated case numbers will be used to assess the ability to remain within health system capacity represented by the clinical pathways model, given current levels of social intervention. Such evidence will support strengthening and, when appropriate, cautious relaxation of distancing measures. Further work will examine the impact of varying the intensity of measures over time, to inform the necessary conditions that would enable ‘exit strategies’ from current stringent lockdown conditions in order to ensure maintenance of social and economic functioning over an extended duration.

All of these strategies, which combine to ‘flatten the curve’, will buy time for further health system strengthening and sourcing of needed supplies. Protecting the health and wellbeing of health care workers will be essential to ensure ongoing service provision. ICU capacity will need to be increased several-fold in anticipation of the looming rise in cases.

There are multiple challenges to be overcome along the pathway to delivery of safe and effective COVID-19 vaccines and timeframe to availability is highly uncertain (19). The search for effective therapies continues. Reducing COVID-19 morbidity and mortality therefore relies on broadly applied public health measures to interrupt overall transmission, protection of vulnerable groups, and ongoing attention to maintaining and strengthening the capacity of health care systems and workers to manage cases.

## Data Availability

Not applicable - modelling study

---

**Supplementary Figure 1:**
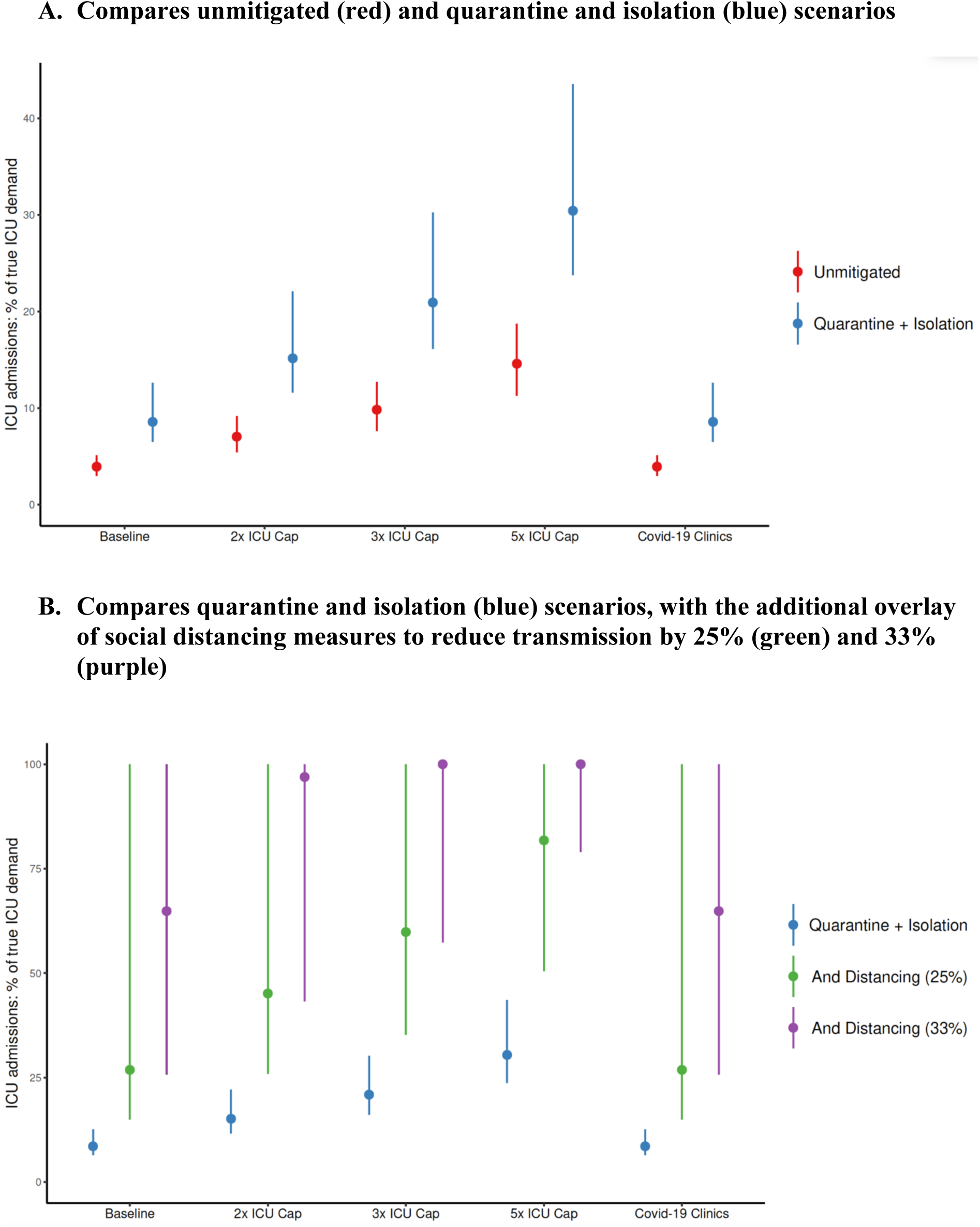
Total ICU admissions throughout the course of the epidemic, as a percentage of true critical care demand given baseline, 2, 3 and 5 × ICU capacity for COVID-19 admissions. The ‘COVID-19 clinics’ scenario reflects an alternative triage pathway, and baseline capacity. Dots denote medians, lines range from 5^th^ to 95^th^ centiles of simulations.

**Supplementary Figure 2:**
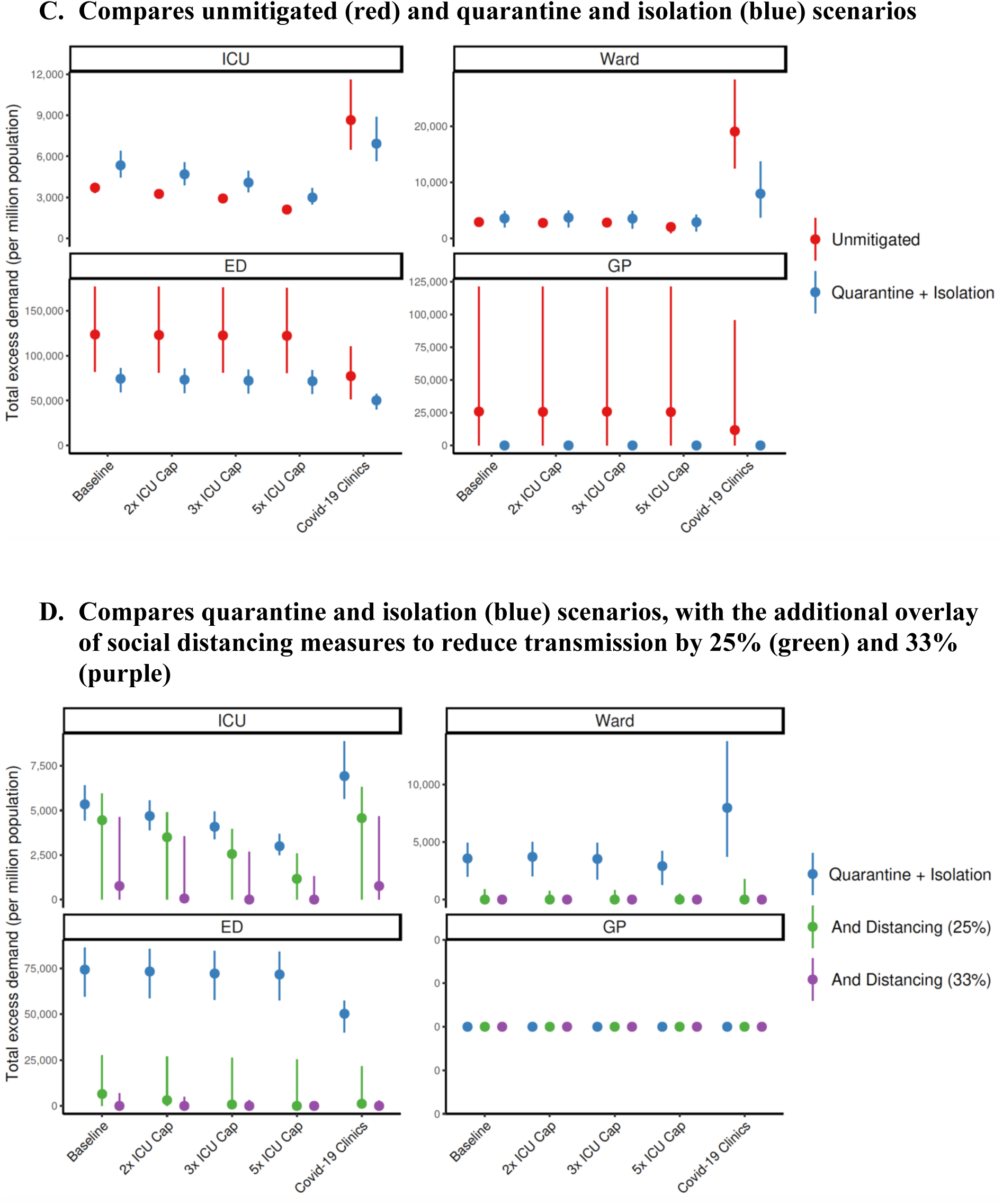
Total excess demand for services assessed by standard care pathways given baseline, 2, 3 and 5 × ICU capacity for COVID-19 admissions, or the alternative triage pathway (with baseline capacity). Dots denote medians, lines range from 5^th^ to 95^th^ centiles of simulations.

## References

1. Coronavirus disease 2019 (COVID-19) Situation Report - 75. 4 April 2020, World Health Organisation.

2. Wu Z, McGoogan JM. Characteristics of and Important Lessons From the Coronavirus Disease 2019 (COVID-19) Outbreak in China: Summary of a Report of 72314 Cases From the Chinese Center for Disease Control and Prevention. JAMA. 2020.

3. Chen N, Zhou M, Dong X, Qu J, Gong F, Han Y, et al. Epidemiological and clinical characteristics of 99 cases of 2019 novel coronavirus pneumonia in Wuhan, China: a descriptive study. Lancet. 2020;395(10223):507–13.

4. Remuzzi A, Remuzzi G. COVID-19 and Italy: what next? Lancet. 2020.

5. Emanuel EJ, Persad G, Upshur R, Thome B, Parker M, Glickman A, et al. Fair Allocation of Scarce Medical Resources in the Time of Covid-19. N Engl J Med. 2020.

6. Australian Health Management Plan for Pandemic Influenza. Australian Government Department of Health, 2019.

7. Australian Health Sector Emergency Response Plan for Novel Coronavirus (COVID-19). Australian Government Department of Health, 2020.

8. McCaw JM, Glass K, Mercer GN, McVernon J. Pandemic controllability: a concept to guide a proportionate and flexible operational response to future influenza pandemics. J Public Health (Oxf). 2014;36(1):5–12.

9. Coronavirus (COVID-19) current situation and case numbers. Australian Government Department of Health [Available from: https://www.health.gov.au/news/health-alerts/novel-coronavirus-2019-ncov-health-alert/coronavirus-covid-19-current-situation-and-case-numbers.

10. Moss R, McCaw JM, Cheng AC, Hurt AC, McVernon J. Reducing disease burden in an influenza pandemic by targeted delivery of neuraminidase inhibitors: mathematical models in the Australian context. BMC Infect Dis. 2016;16(1):552.

11. Lai S, Ruktanonchai N, Zhou L, Prosper O, Luo W, Floyd J, et al. Effect of non-pharmaceutical interventions for containing the COVID-19 outbreak: an observational and modelling study. medRxiv preprint. 2020.

12. Cowling B, Ali S, Ng T, Tsang T, Li J, Fong M, et al. Impact assessment of non-pharmaceutical interventions against COVID-19 and influenza in Hong Kong: an observational study. medRxiv preprint. 2020.

13. Ferguson N, Laydon D, Nedjati-Gilani G, Imai N, Ainslie K, Baguelin M, et al. Impact of non-pharmaceutical interventions (NPIs) to reduce COVID-19 mortality and healthcare demand. Imperial College COVID-19 Response Team. 2020.

14. Walker P, Whittaker C, Watson O, Baguelin M, Ainslie K, Bhatia S, et al. The global impact of COVID-19 and strategies for mitigation and suppression. WHO Collaborating Centre for Infectious Disease Modelling, MRC Centre for Global Infectious Disease Analysis, Abdul Latif Jameel Institute for Disease and Emergency Analytics, Imperial College London; 2020.

15. Zhang J, Zhou L, Yang Y, Peng W, Wang W, Chen X. Therapeutic and triage strategies for 2019 novel coronavirus disease in fever clinics. Lancet Respir Med. 2020;8(3):e11–e2.

16. Cheng AC, Holmes M, Dwyer DE, Senanayake S, Cooley L, Irving LB, et al. Influenza epidemiology in patients admitted to sentinel Australian hospitals in 2017: the Influenza Complications Alert Network (FluCAN). Commun Dis Intell (2018). 2019;43.

17. Legido-Quigley H, Asgari N, Teo YY, Leung GM, Oshitani H, Fukuda K, et al. Are high-performing health systems resilient against the COVID-19 epidemic? Lancet. 2020;395(10227):848–50.

18. Moss R, Fielding JE, Franklin LJ, Stephens N, McVernon J, Dawson P, et al. Epidemic forecasts as a tool for public health: interpretation and (re)calibration. Aust N Z J Public Health. 2018;42(1):69–76.

19. Lurie N, Saville M, Hatchett R, Halton J. Developing Covid-19 Vaccines at Pandemic Speed. N Engl J Med. 2020.

20. Wu JT, Leung K, Leung GM. Nowcasting and forecasting the potential domestic and international spread of the 2019-nCoV outbreak originating in Wuhan, China: a modelling study. Lancet. 2020;395(10225):689–97.

21. Li Q, Guan X, Wu P, Wang X, Zhou L, Tong Y, et al. Early Transmission Dynamics in Wuhan, China, of Novel Coronavirus-Infected Pneumonia. N Engl J Med. 2020;382(13):1199–207.

22. Lauer SA, Grantz KH, Bi Q, Jones FK, Zheng Q, Meredith HR, et al. The Incubation Period of Coronavirus Disease 2019 (COVID-19) From Publicly Reported Confirmed Cases: Estimation and Application. Ann Intern Med. 2020.

23. Ganyani T, Kremer C, Chen D, Torneri A, Faes C, Wallinga J, et al. Estimating the generation interval for COVID-19 based on symptom onset data. medRxiv preprint. 2020.

24. Tindale L, Coombe M, Stockdale J, Garlock E, Lau W, Saraswat M, et al. Transmission interval estimates suggest pre-symptomatic spread of COVID-19. medRxiv preprint. 2020.

25. Report on 775 patients critically ill with COVID-19. Intensive care national audit and research centre. [Available from: https://www.icnarc.org/About/Latest-News/2020/03/27/Report-On-775-Patients-Critically-Ill-With-Covid-19.

26. Epidemia COVID-19. Aggiornamento nazionale.. 26 marzo 2020.

